# A pipeline for tabular dataset formation from unstructured data provided by ACR Appropriateness Criteria guidelines

**DOI:** 10.1101/2022.04.20.22274096

**Authors:** Anderson A. Eduardo, Rafael M. Loureiro, Adriano Tachibana, Pedro V. Netto, Tatiana F. de Almeida, Luiz Henrique Alves Monteiro, André P. dos Santos

## Abstract

Currently, data performns a critical concept for disparate human activities, from law to technology. Among data-centric technologies, clinical decision support systems (CDSS) figures out as one of the most promising for healthcare. Despite the technological advances facilitating its implementation, the maintainance of knowledge base for CDSS remains open to improvements. Here, we argue that the Appropriateness Criteria provided by ACR guidelines can be used as a open data-source that, combined with appropriate algorithms, can push forward basic research and technological developments regarding knowledge base for CDSS. Therefore, we developed a pipeline capable of forming tabular datasets from ACR guidelines, stored in a web site as textual `PDF` files. We also experimentally demonstrate that the proposed pipeline successfully recorvers the interested contents, and the best composition, in terms of its component algorithms, is discussed. Future research focused on algorithms flexibility in the face of `PDF` template updates could improve our work.

## 1 Introduction

In the last decade, the concept of data has become a central topic for a wide range of human activities. Concerns on how data is collected, its contents, where and how it is stored, how it could be accessed, among other issues, currently permeate even disparate intellectual and practical areas of inquiry, from law to mathematics, passing through virtually all fields related to modern digital technology. For health sciences, the historical importance of data collection and its availability dramatically deepened with the dawn of data-centric paradigm [10].

Among current data-centric technologies in health sciences, decision support systems (and, more specifically, clinical decision support systems - CDSS for short) figures out as one of the most promising concept [8, 5, 9, 1]. Practically speaking, a CDSS is implemented as a software, usually integrated with previously existing information systems [9]. Such software runs under the rotinetely use of a local system, displaying some type of signalization to the user when the prescription of clinical exams is detected. The signalization delivers sugestions of best practices, given patient information [9].

One of the most critical step in implementing a CDSS regards the knowledge base formation and maintainance. In the first deployed systems, this work was done manually and was highly labor-intensive. But in the recent years, very flexible and performatic data structures becomes ready to use, greatly facilitating the implementation of knowledge bases. Even so, the maintainance remains open to advances.Guidelines for professional practice presents a rich and effective source of knowledge for clinical decision support systems. Specifically for radiology, the American College of Radiology (hereafter, ACR) maintains the Appropriateness Criteria guidelines, a web resource in which a team of experts provide a series of high-quality guidance for prescription of radiological exams. The ACR guidelines are presented in Portable Document Format (*i*.*e*., `PDF` files), listed in a searchable webpage, organized by clinical specialities. The access is free and open, just navigate to the `URL` https://acsearch.acr.org/list. Such resource encompasses the very nature of a knowledge base for CDSS.

Naturally, the ACR guidelines should be considered a major information source for clinical decision support in radiology (and, in fact, it is). However, it is designed to human readers, being presented in unstructured, textual `PDF` files, not easily prone to machine reading. We argue that ACR guidelines are too valuable data-resource and computational algorithms and pipelines able to parse it to structured data could help to push forward both basic research and application developments regarding CDSS for radiology.

Thus, in the present work, we conceptualize a pipeline for processing ACR guidelines to a structured tabular dataset. We also provide a `python` implementation, as well as a benchmark experiment, where correctness and computational performance were assessed.

## 2 Background

Currently, research on the different aspects of data collection by computational means unfolded in a number of research programs, each one focused on slightly different aspects of data collection, its levels of complexity and types of digital applications. Inevitably, the literature is huge, spanning different areas of computer science for decades. Examples are text mining, web mining, document understanding, and information retrieval, just for naming some of them [11, 12, 6, 7, 3]. Despite of theoretical and technical specificities, raw data must be collected (or recovered) at some step.

In the context of digital health science projects, Towbin (2019) [10] points out that data collection, besides data analysis, is a core activity, specially in radiology. Data collection can be performed through different approaches. In manual data collection, a few people (or even only one) conduct all the work. It also can be carried out in a distributed manner, by professionals in disparate organizations or geographical locations. This last approach is currently employed by a number of research teams in a world-wide scale [10]. The main advantage concerns that the collection itself probably could be done by any minimally trained person and, in the case of multi-institutional teams, data could be available in a short time-frame. The main disadvantage is related to data quality and heterogeneity, as raw data potentially has been collected using different protocols and equipment. Moreover, the manually collected data is naturally prone to human error [2].

Eletronic data collection is the most used approach, as digital storage of large volumes of data has become cheaper over the last few decades and database management systems have advanced to become more palatable to users [4]. The advantage of this approach regards the large volume and accessability provided by current data-base applications. Also, such applications can be moderately authomated, increasing the availability of relevant data. But specialized professionals are demmanded to structure the data-base application, specially in cases of data ingestion from multiple sources.

Fully authomated data collection, extraction and evaluation is currently possible, thanks to advances in artificial intelligence algorithms and computational power [10]. By this approach, disparate and unstructured data can be processed along with structured data. The final datasets can be presented in tabular data structures, convenient for data analytics and machine learning. This approach has tremendously impacted digital data collection [10]. The down side is that highly specialized professionals are demanded and a longer time-frame is needed till the application for data collection is up to be used.

A number of conceptual and practical developments has been done for electronic data collection and fully automated data collection. Despite of that, we were not able to find in the literature any work that has specifically focused on the formation of structured datasets from the ACR guidelines. Being Appropriateness Criteria provided by ACR a valuable data-source, we proceeded with a first conceptual approach and computational implementation and experimentation for tabular dataset formation from such data-source.

## 3 The proposed pipeline

The `PDF` documents of ACR guidelines are relatively well structured texts, presenting its main contents (which are clinical indications and its respective recommendations of radiological exams) in tables and in visually distinguishable locations throughout the documents. Despite of that, a number of other tables and textual contents usually are presented, such as expert considerations, synthesis of available empirical data, and referencial literature. More over, the interested contents usually appear in many different places, composing different document sections. An example can be viwed at figure (1).

**Figure 1:**
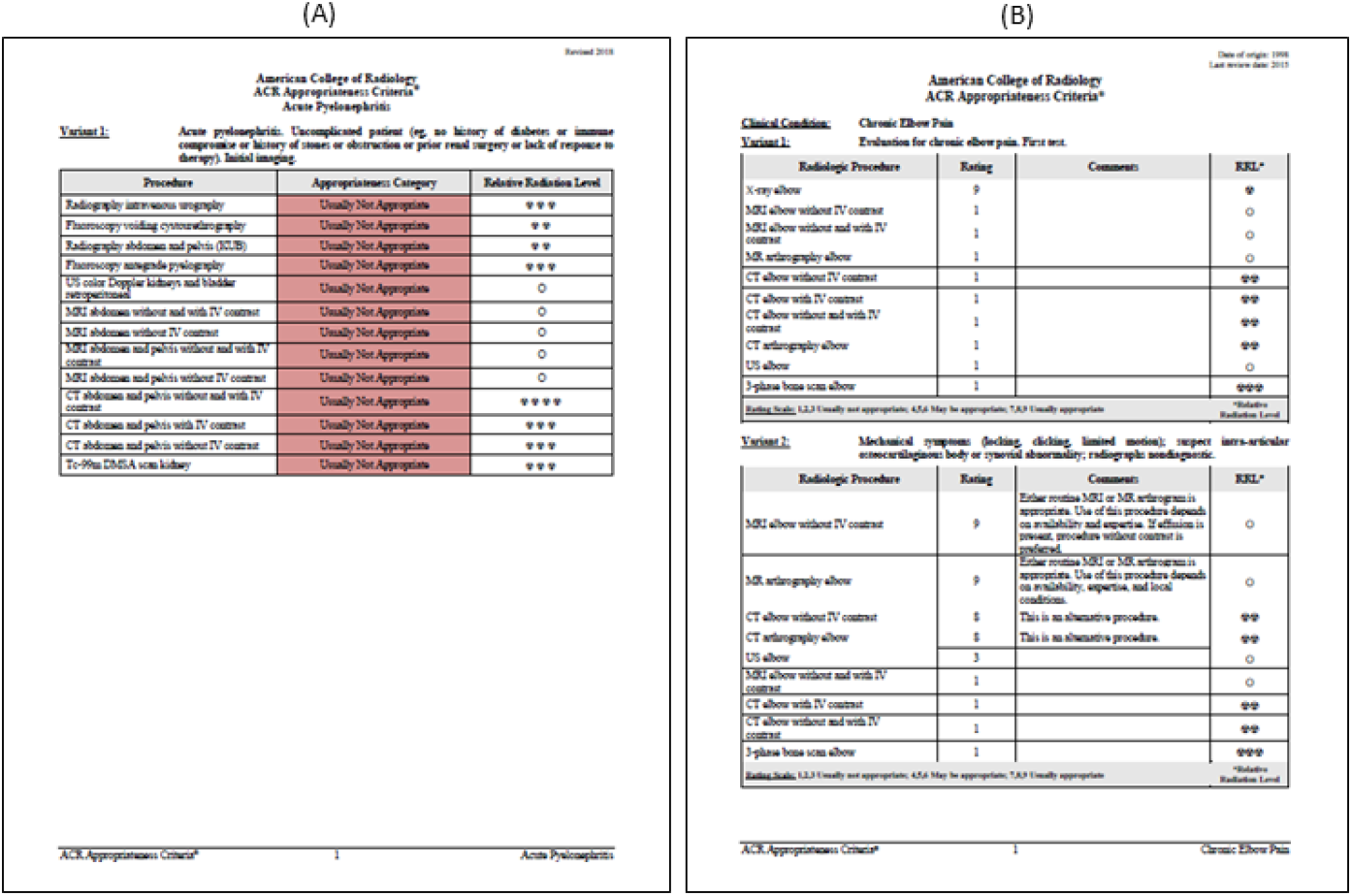
Examples of PDF files provided by ACR guidelines. Note that slightly different templates are used by ACR.

**Figure 2:**
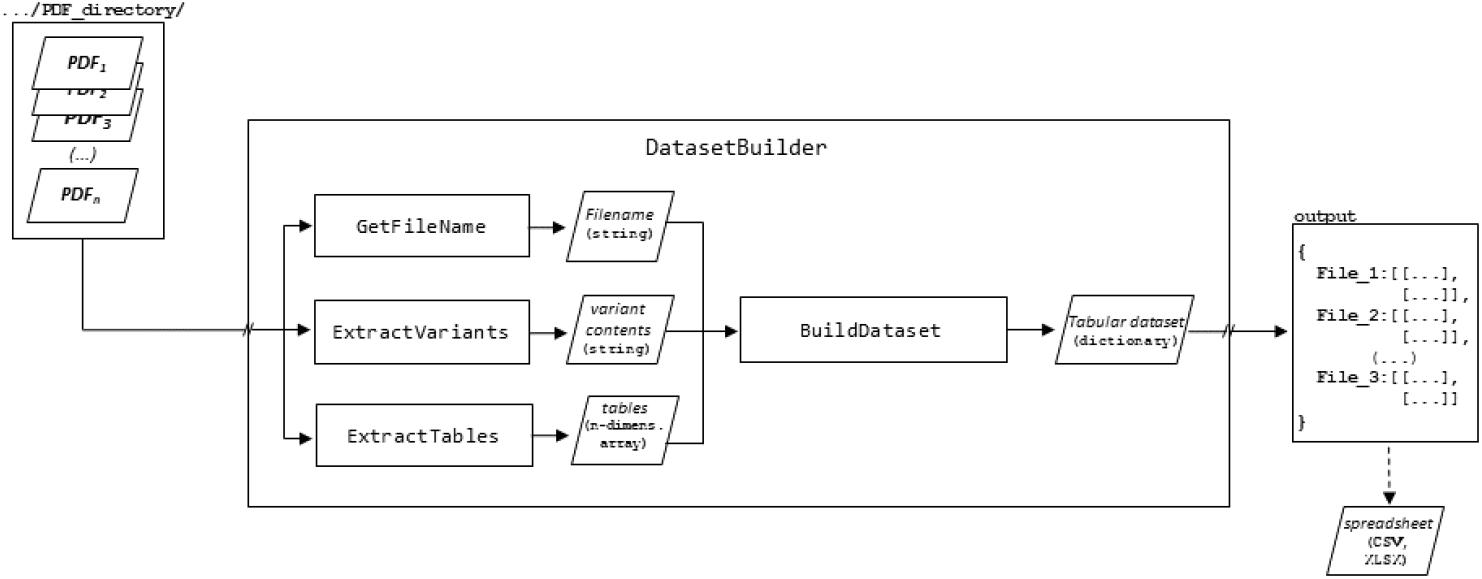
Diagrammatic representation of the proposed pipeline, showing the component algorithms and indication of data structures.

**Figure 3:**
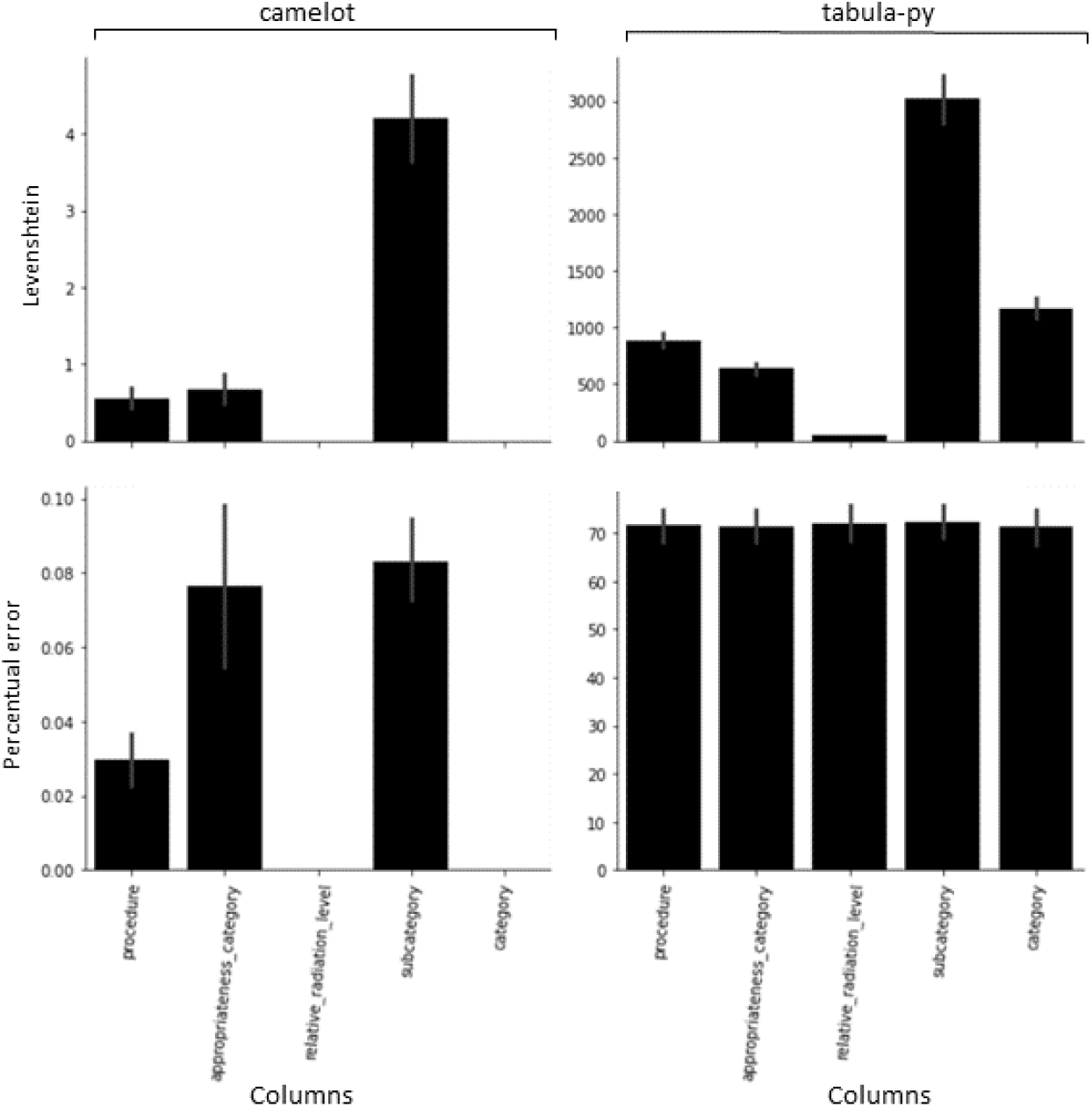
Experiment results for comparison between output datasets, produced using the proposed pipeline, and the benchmark dataset. All comparisons were performed in terms of whole column contents. The percentual error is computed dividing the Levenshtein value by the total number of characters in the reference dataset (details in the text). Note that y-axes are in different scales.

**Figure 4:**
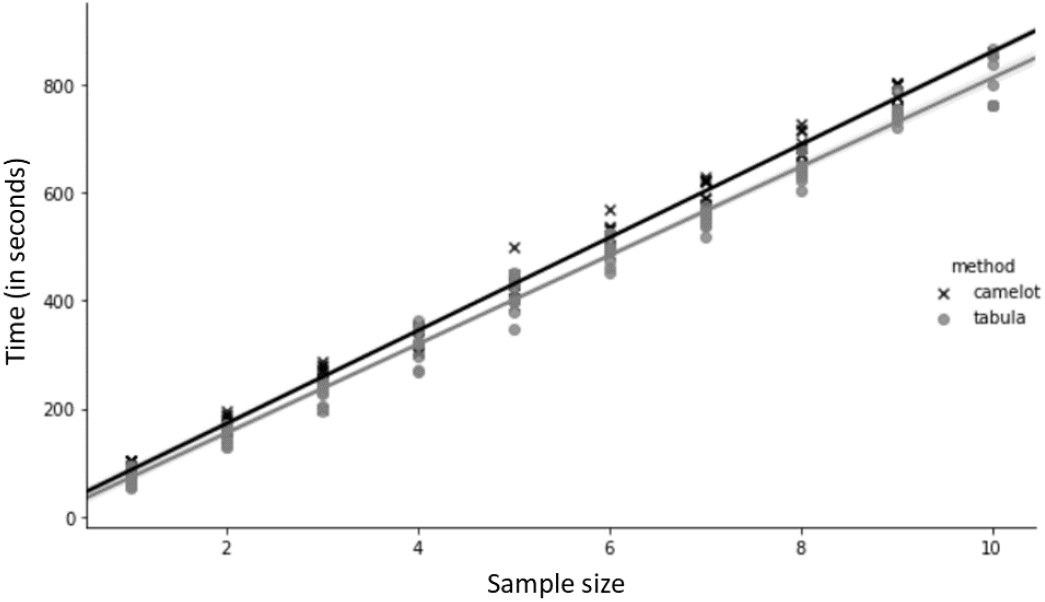
Results of time complexity for the proposed pipeline. The two core functions (*i*.*e*., *tabula-py* and *camelot*) for table extraction are compared. A very similar, linear pattern is observed for both versions of the pipeline.

Here, our contents of interest comprises the *Variant* texts and the tables for *Appropriateness Criteria* (see figure (1)). Also, the filename presents the name of the guideline specific group, providing another piece of interested data for our purposes. Thus, our main algorithm, the `DatasetBuilder` algorithm, is built uppon subalgorithms able to maping specific contents.

At the core of `DatasetBuilder` algorithm, the subalgorithms `GetFileName`, `ExtractVariants`, `ExtractTables` and `BuildDataset` performs the main tasks independently. As the main algorithm iterates over the set of `PDF` filepaths provided by user, the `GetFileName` just manipulate such `string` in order to return the filename from an inputted filepath. The `ExtractVariants` algorithm is an `OCR`-based algorithm which converts each page in the `PDF` file into images, compute all text blocks coodinates throughout the document, performs the convertion from image to text (*i*.*e*., the OCR-procedure itself) and, from the obtained set of `strings`, find *Variant* text blocks, returning it as its output. The `ExtractTables` algorithm is a wrapper for the `tabula-py` and `camelot` python libraries. Both libraries present the computational function `read_pdf()`, which parses file contents and returns the found tables. For a detailed description of the algorithms underling these functions, please refer to the respective projects documentation (https://tabula-py.readthedocs.io/en/latest/index.html, for `tabula-py`; https://camelot-py.readthedocs.io/en/master/ for `camelot`). In a fourth step, the `BuildDataset` receives the output objects from the previous three algorithms and operates on them, in order to restructure such data structures into a single one (specifically, a matrix-like data structure). In tandem, these algorithms are able to extract the interested data from an ACR guideline `PDF` file, returning it as a single, structured tabular data for that file. Finally, the final output from `DatasetBuilder` is a `dictionary`, in which the keys are filenames and the values are tabular data structures bearing the interested contents for each respective file. A diagrammatic representation of our pipeline is shown in figure (1).

## 4 Experiment

In order to accomplish the evaluation of the proposed pipeline, our first step was to produce a reference dataset for benchmark. Thus, we ramdomly selected 10 `PDF` files at ACR Appropriateness Criteria website (https://acsearch.acr.org/list). The interested contents were manually parsed to a common `CSV` file and located in a convenient directory in our project file system.

Our benchmark algorithm iterates over each column name found in the benchmark dataset. This is necessary to constrain our analysis focused in the interested contents, teasing apart possible failures regarding other ACR guidelines content, beyond the scope of our research. Future work should be carried out on this topic. For the selected columns, all of its contents were concatenated into a single textual data structure. Thus, as the proposed pipeline outputs all the interested contents of an ACR guidelines`PDF` file agregated into a single tabular data structure, our benchmark algorithm performs the comparison of its contents by the means of comparing each `string`, which agregates the contents for each respective column, between the dataset produced by the pipeline and the benchmark dataset. For the measurements of the level of match, the Levenshtein distance were used. Also, we compute an percentual error, by dividing the Levenshtein metric value by the total number of characters in the reference `string` (for each column). The values were registered along with `PDF` filename, column name and number of characters in the reference `string` for that column.

To consistently evaluate the proposed pipeline, the benchmark algorithm were structurally encapsulated into a iterative algorithm. This algorithm is responsible for running experiment replicates. This replicates were designed to make it possible to evaluate the time complexity of the proposed pipeline, the performance with different combinations of `PDF` files, the table extraction core methods (*i*.*e*., `tabula-py` and `camelot`), and the stability of the computational implementation. All data referring to iterations were also registered along with the metric value, as described in the paragraph above. The Algorithm (1) (`BenchmarkExperiment`) provides the pseudocode representation for our benchmark algorithm.

### Algorithm 1 The *BenchmarkExperiment* algorithm

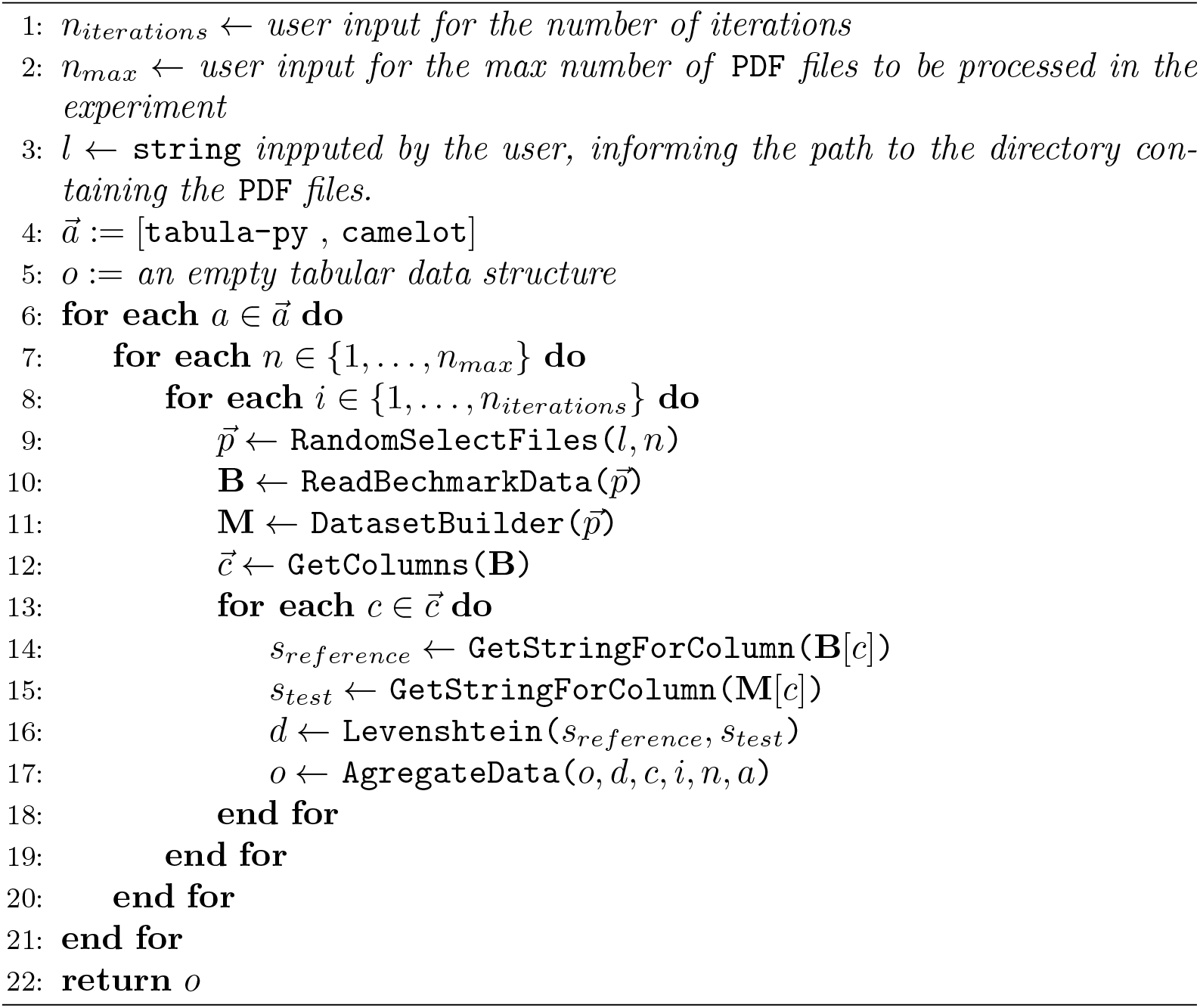

All the implementations were conduced using `python` version 3.8.8, in a personal computer with an `Intel(R) Core(TM) i5-10210U`, 1.60*GHz−*2.11*GHz* `CPU` and 16*GB* `RAM`. The data produced by the experiment was analyzed graphically, using `Matplotlib` version 3.3.4 and `Seaborn`version 0.11.1. All the implementation code is provided via a dockerized project, available at https://github.com/AndersonEduardo/pipeline_acr_guidelines.

## 5 Results and Discussion

Experimental results show that the proposed pipeline was able to recover up to 100% of our benchmark dataset for the columns *Relative Radiation Level* and *Category*. Also, a high performance was observed for the other columns, specially *Procedure* and *Appropriateness Category*. Strictly speaking, the lower performance was observed for *Subcategory* (Levenshtein distance of 4), but it must be noted that the percentual error was *<* 0.10% (figure (5)).

The core function employed in the `ExtractTables` algorithm strongly affected the pipeline output. The best performance was observed only when `camelot` is employed. Using `tabula-py`, the whole performance decays to critical levels, with the interested contents being only loosely recovered. In some cases, whole tables were not recovered, compromising the final pipeline output. In fact, `camelot` is built uppon `tabula-py`, improving many of its algorithms. Despite of that, the performance results for `tabula-py` do not resembles the one observed for `camelot`, meaning that the relateness between these `python` libraries is not translated in terms of similar performance for our pipeline. For the range of input files considered in our experiment, the empirical time complexity shows a linear pattern (figure (5)). Moreover, it was very similar for both `camelot` and `tabula-py`, being only sensitively lower for the second one. We attribute such observations to the fact that `tabula-py` loses some tables, thus it is prone to parse slightly less data from de `PDF` files. In other words, the better performance provided by `camelot` does not take additional time cost, in relation to `tabula-py`. Future work should be carried out on this topic, in order to explore a wider range of input-files number (ideally, in terms of hundreds of `PDF` files).

## 6 Conclusion

In this work, we proposed an experimental pipeline designed to form tabular datasets from online Appropriateness Criteria guidelines of ACR. Combining authoral and third party algorithms (open source), our proposed pipeline relies on NLP and computer vision concepts and technics, being able to successfully parse `PDF` files from ACR guide-lines. Taken together, experimental results shown that our approach recovered the contents of our benchmark dataset with an percentual error of *<* 0.1%, when `camelot` is employed for table extraction. Through the pipeline, the Appropriateness Criteria data from ACR becomes readily accessible for machine learning and data analytics studies. The `python` implementation is available and it shows a good computational performance even in a ordinary personal computer.

Future work should focus on making the pipeline algorithms more flexible, in view of stability in the face of changes or updates to `PDF` templates by ACR.

## Data Availability

All data produced in the present study are available upon reasonable request to the authors.

https://acsearch.acr.org/list

